# The Interoceptive–Metacognitive Architecture of Fatigue: Partial Mediation and Resilience Buffering

**DOI:** 10.1101/2025.09.12.25335659

**Authors:** Vitalii Lunov, Bohdan Tkach, Mykhailo Matiash, Natalia Yevdokymova

**Affiliations:** Bogomolets national medical university; Pylyp Orlyk International Classical University; Bogomolets National Medical University

**Keywords:** interoception, predictive processing, metacognition, fatigue, psychosocial functioning, academic resilience

## Abstract

**Background:** The problem here to be studied is whether interoceptive predictive processing and metacognitive appraisal jointly explain persistent fatigue and everyday impairment. Contemporary theory suggests that sustained interoceptive prediction error undermines allostatic self-efficacy, with fatigue operating as a metacognitive signal that expected costs outweigh benefits.

**Methods:** We conducted a preregistered, cross-sectional study in a UA/EU higher-education cohort (N = 300, age 18–35). Participants completed MAIA-2 (interoceptive sensibility), MCQ-30 (metacognitive beliefs), BIS/BAS (approach– avoidance), FSS-9 (fatigue), IPF (psychosocial functioning; coded higher = greater impairment), and ARS-30 (academic resilience). An ESEM measurement model (with multi-group invariance by sex and region) preceded a structural model estimating direct paths to IPF and an indirect path via FSS; ARS-30 was tested as a moderator. Robust maximum likelihood was used with FIML for missing data; indirect effects employed bootstrap CIs.

**Results:** A three-factor ESEM (Interoceptive Sensibility, Metacognitive Beliefs, Approach–Avoidance) fit well and showed metric—plus partial scalar— invariance (sex, UA/EU). In structural analyses, metacognitive threat/uncontrollability was associated with higher fatigue and greater impairment, whereas interoceptive sensibility showed protective associations (lower fatigue, better functioning). Fatigue carried a substantive portion of metacognitive influence on impairment (partial mediation), with residual direct effects intact (complementary mediation). Approach–avoidance tendencies contributed modestly. Academic resilience attenuated the metacognitive threat → fatigue pathway. Core paths were invariant across sex and region.

**Conclusions:** Findings are consonant with predictive-processing and cost– benefit control accounts: persistent expectation–evidence mismatch amplifies perceived costs and degrades confidence in bodily control, with fatigue as the gatekeeper to down-regulated action. The work establishes a measurement-tight platform for longitudinal and post-exertional paradigms incorporating behavioural interoceptive accuracy/insight to quantify prediction error directly.

## Introduction

Fatigue is a ubiquitous experience in both health and disease, yet its mechanisms remain contested. The problem here to be studied is whether disruptions in interoceptive predictive processing—specifically the persistence of mismatches between prior expectations and ascending bodily evidence—together with metacognitive appraisal of controllability, can account for the transition from transient tiredness to persistent, function-limiting fatigue. Contemporary accounts grounded in predictive processing and allostatic self-efficacy propose that sustained interoceptive prediction error degrades confidence in the brain’s internal control models; fatigue is then experienced as a metacognitive signal that expected costs of action outweigh expected benefits (Stephan et al., 2016; Greenhouse-Tucknott et al., 2021). Thus, the question therefore naturally arises as to how interoceptive inference and metacognitive beliefs jointly shape day-to-day functioning.

A converging empirical literature lends this framework credibility while exposing important gaps. Clinical and experimental studies indicate that facets of interoception dissociate: patients with fibromyalgia, ME/CFS or MS may show intact objective accuracy but elevated interoceptive “prediction error” (discrepancy between subjective sensibility and objective accuracy) that tracks pain and fatigue (Sharp et al., 2021; Campo et al., 2019). In MS, lower interoceptive insight—together with white-matter dysconnectivity—elevates the odds of cognitive fatigue, whereas questionnaire-based interoceptive awareness and sleep quality prospectively predict individual fatigue scores (Danciut et al., 2024; Rouault et al., 2023). Functional imaging further implicates anterior insula and dorsal anterior cingulate—regions central to interoceptive integration and error monitoring—with altered connectivity to reward circuitry during fatigue states (Chen et al., 2020). These findings align with a broader neurobiological synthesis that situates interoception as the afferent arm of homeostatic and allostatic control (Quadt et al., 2018).

Another way of looking at this question is to embed predictive coding within a cost–benefit controller. Neurocognitive models argue that the anterior cingulate cortex integrates error, effort and reward, with catecholaminergic systems energising goal pursuit; when predicted costs chronically exceed expected benefits, behaviour is down-regulated and fatigue is reported (McMorris et al., 2018; McMorris, 2020; Kok, 2022). Task-based evidence shows that the very cingulate territories recruited by error processing co-vary with on-task fatigue (Wylie et al., 2017). At the computational level, trial-wise modelling demonstrates that momentary fatigue updates as a function of exerted effort (and errors in the cognitive domain) and prospectively alters subsequent effort choices, linking inference to behaviour in real time (Matthews et al., 2023). Learning accounts add that interoceptive and exteroceptive cues can acquire predictive value for “feeling tired,” thereby fostering avoidance and generalisation that entrench disability (Lenaert et al., 2018). Teodoro et al. (2018) further suggest that excessive interoceptive monitoring and heightened perceived effort reallocate attention inward, slowing processing and amplifying the felt cost of routine actions across fatigue-laden conditions.

Despite these advances, two gaps remain prominent. First, few studies integrate interoceptive sensibility (as a measurable, multidimensional construct), metacognitive beliefs about uncontrollability and danger, approach–avoidance tendencies, and resilience within a single structural model that treats psychosocial functioning as the criterion outcome and fatigue as a putative mediator. Second, measurement rigour—specifically, defensible latent structures and tests of measurement invariance across populations—has not been consistently demonstrated outside disorder-defined cohorts. The present study addresses these gaps in a UA/EU higher-education sample by combining a pre-registered ESEM–SEM pipeline with harmonised reporting and an explicit outcome direction (higher IPF scores denote greater impairment).

## THEORETICAL FRAMEWORK

The problem here to be studied is the apparent convergence between theories of interoceptive predictive processing and the lived, disabling experience of pathological fatigue. The question therefore naturally arises as to how disruptions in interoceptive inference—specifically persistent prediction error and diminished metacognitive confidence in bodily control—scale up from momentary effort costs to prolonged states of fatigue that characterise clinical conditions such as ME/CFS, fibromyalgia, multiple sclerosis (MS) and major depressive disorder (Greenhouse-Tucknott et al., 2021; Stephan et al., 2016; Quadt et al., 2018; Rouault et al., 2023; Campo et al., 2019; Eggart et al., 2023). At the heart of the discussion is the claim that the brain’s generative model of bodily states—subserved by insula–cingulate–prefrontal circuits and modulated by catecholaminergic systems—computes cost–benefit predictions that integrate interoceptive evidence, motivational value and control capacity; when these computations chronically fail to resolve mismatch, fatigue ensues as a metacognitive diagnosis of low allostatic self-efficacy (Stephan et al., 2016; McMorris et al., 2018; McMorris, 2020; Kok, 2022).

The doctrine provided by predictive processing frames interoception as hierarchical inference: top-down priors about internal milieu are compared with ascending viscerosensory evidence, yielding prediction errors that tune beliefs and actions. We would like to dwell on the problem of how such errors become persistent and phenomenologically “sticky”. Greenhouse-Tucknott and colleagues (2021) explicitly unify exertional and pathological fatigue, arguing that sustained detection of interoceptive mismatch undermines confidence in control predictions; fatigue is then the conscious signal that forecasted effort– benefit trade-offs have turned unfavourable. This hypothesis is in accord with the facts established by work in MS and chronic pain/fatigue syndromes, where facets of interoception—accuracy, sensibility, and metacognitive insight—are dissociable and differentially linked to symptom burden (Rouault et al., 2023; Sharp et al., 2021; Danciut et al., 2024). Specifically, patients may exhibit intact accuracy yet elevated “interoceptive prediction error” (the discrepancy between subjective sensibility and objective accuracy), and this trait correlates with fatigue, pain and reduced pain thresholds (Sharp et al., 2021). Danciut et al. (2024) further show that low interoceptive insight—and its interaction with white-matter dysconnectivity—predicts cognitive fatigue in MS, thereby grounding metacognitive failure in measurable structural constraints.

Another way of looking at this question is to bring motivational and control architectures into the same frame. McMorris and colleagues (2018; 2020) propose that dorsolateral prefrontal cortex (DLPFC) generates predictions of effort and sensory consequences that are broadcast to anterior insula (AIC) and cingulate cortex; the dorsal ACC (dACC) links effort costs to reward valuation, with dopaminergic (VTA/SNc) and noradrenergic (LC) systems energising goal pursuit. Kok (2022) similarly argues for a cost–benefit controller in medial prefrontal cortex that integrates fatigue signals into decisions about continued exertion. We must therefore hold that fatigue is not merely a passive by-product of depletion; rather, it is a decision-relevant signal that down-weights the net value of action when predicted costs repeatedly exceed expected benefits. Converging evidence from task fMRI shows that the very ACC territories engaged by error monitoring and outcome prediction co-vary with on-task fatigue, consistent with a shared computational substrate for error processing, effort valuation and fatigue signalling (Wylie et al., 2017). Matthews et al. (2023) sharpen this claim by demonstrating trial-by-trial dynamics: momentary fatigue increases as a function of exerted effort (and errors in the cognitive domain) and prospectively alters subsequent effort choices—precisely the signature expected under an inference-based controller.

The subject of the investigation is, thus, the interplay between interoceptive inference and learning. The ALT+F model (Lenaert et al., 2018) formalises how interoceptive and exteroceptive cues can become associated with fatigue via associative learning, promoting avoidance and generalisation that entrench symptom persistence. The theory was applied to interpret how innocuous bodily cues acquire predictive value for “feeling tired,” thereby biasing priors towards high expected effort and low expected control. This dovetails with empirical neuroimaging in MS showing hyperconnectivity among interoceptive hubs (insula, dACC) and relative disconnection within reward circuitry during fatigue states (Chen et al., 2020; Campo et al., 2019), again supporting a model in which aberrant precision-weighting of interoceptive signals and degraded valuation jointly amplify the subjective cost landscape.

The problem of post-exertional worsening (PEW/PEM) is in the focus of attention of current debates because it provides a naturalistic perturbation to test predictive accounts. The most common hypothesis in this niche is that an exertional challenge temporarily elevates interoceptive prediction error and shifts motivational priors, producing delayed and sometimes disproportionate fatigue responses (Greenhouse-Tucknott et al., 2021; Matthews et al., 2023). While the precise kinetics likely vary across disorders, the logic—persistent error → reduced confidence in control → down-regulated action policies—has explanatory reach across physical and cognitive domains. It is a mistaken view to treat interoception as a monolith: the weight of evidence indicates that “insight” (metacognitive calibration of one’s accuracy) is a stronger determinant of fatigue than raw accuracy per se in at least some cohorts (Danciut et al., 2024; Rouault et al., 2023). We have come to accept that this metacognitive layer—captured by Stephan et al.’s (2016) allostatic self-efficacy—links dyshomeostasis to affective and motivational sequelae, including depression, thereby offering a principled account of comorbidity patterns.

Finally it can be observed that this synthesis does not dismiss alternative mechanisms—neuroinflammation, monoaminergic tone, sleep disruption— but reinterprets them as modulators of precision and controllability within an inferential controller (Quadt et al., 2018; Kok, 2022; Eggart et al., 2023). From this we can conclude that a viable path forward is to test interventions that specifically target (i) precision-recalibration of interoceptive signals (e.g., biofeedback or tasks that reduce sensibility–accuracy discrepancy), (ii) enhancement of interoceptive insight (metacognitive training anchored in objective feedback), and (iii) cost–benefit reweighting of effort valuation (motivational framing, dopaminergic/adrenergic modulation where appropriate). The idea still needs considerable working out in prospective and disorder-defined cohorts, but the theoretical scaffolding now ties together what used to be separate literatures on exertional, cognitive and pathological fatigue (Greenhouse-Tucknott et al., 2021; Matthews et al., 2023; Stephan et al., 2016; McMorris, 2020).

## Materials and Methods

We conducted a cross-sectional, questionnaire-based study with a pre-specified modelling pipeline: (i) measurement modelling via exploratory structural equation modelling (ESEM) for latent predictors; (ii) multi-group tests of measurement invariance by sex and region (UA vs EU); and (iii) a structural model estimating direct paths to Impairment in Psychosocial Functioning (IPF) and an indirect path via fatigue (FSS-9). The analysis plan, codebook, and scripts were preregistered prior to data lock.

To preserve ecological validity for academic resilience while ensuring variability in interoceptive and fatigue indices, we recruited students enrolled in higher-education programmes.

We enrolled N = 300 adults aged 18–35 years, approximately evenly split between Ukraine (n≈150) and continental EU (n≈150), with near-balanced sex representation. Inclusion criteria were: age 18–35, current enrolment (full-or part-time) in higher education, proficiency in Ukrainian **or** English sufficient for survey completion, internet access, and informed consent. Exclusion criteria were self-reported current psychosis, neurodegenerative disease, uncontrolled endocrine disease, acute febrile illness, or inability to consent. Self-reported medical diagnoses (e.g., ME/CFS, MS, MDD) were recorded for descriptive purposes only and did not determine eligibility.

Participants were recruited via faculty mailing lists and virtual learning environments. All provided electronic consent and could enter a non-coercive prize draw.

### Procedure

After consent, participants completed the battery online (Qualtrics) in a fixed order to minimise context effects: demographics → MAIA-2 (validated Ukrainian version where available; otherwise English) → MCQ-30 → BIS/BAS → FSS-9 → IPF → ARS-30. Two attention-check items were embedded. Median completion time was 25–35 minutes. A debriefing page and support contacts were provided upon submission.

### Measures

We assessed the following constructs using established instruments and harmonised scoring conventions.

Interoceptive sensibility — MAIA-2 (2018). We analysed theoretically relevant subscales (e.g., Noticing, Attention Regulation, Body Listening, Body Trusting, Not-Worrying). Subscale item-means were used; higher scores indicated greater interoceptive sensibility/skills.

Metacognitive beliefs — MCQ-30. We computed item-means (1–4) for Positive Beliefs about Worry, Negative Beliefs about Uncontrollability and Danger (Threat/Uncontrollability), Cognitive Confidence, Need to Control Thoughts, and Cognitive Self-Consciousness. Higher values indicated stronger endorsement of the belief domain.

Approach–avoidance motivation — Carver & White’s BIS/BAS. We used subscale means for BIS and BAS (Drive, Fun Seeking, Reward Responsiveness); higher values reflected stronger trait tendencies.

Fatigue — FSS-9. The total score (9–63) served as the primary fatigue index and the mediator in the indirect pathway (higher = more severe fatigue).

Psychosocial functioning (primary outcome) — IPF. We explicitly coded IPF such that higher scores denoted greater impairment. A latent IPF factor was formed from four broadly applicable domains (Work/Study, Self-Care, Family/Intimate, Social/Friendships), modelled as continuous parcels.

Academic resilience (moderator) — ARS-30. The total score indexed academic resilience and was tested as a moderator of Threat/Uncontrollability → FSS and Threat/Uncontrollability → IPF paths. In sensitivity analyses, ARS-30 subscales were optionally specified as indicators of a latent resilience factor.

### Data management and quality control

Data were stored on encrypted university servers. Protocol-defined exclusions removed records with >20% missing data. Remaining item-level missingness was handled under full information maximum likelihood (FIML), assuming missing at random. We screened for straight-lining, failed attention checks, and implausibly short completion times.

#### Statistical analysis

### Measurement modelling

Predictor domains (MAIA-2, MCQ-30, BIS/BAS) were specified as three factors and estimated using ESEM with target rotation to permit small cross-loadings. Reliability was indexed with McDonald’s ωat the subscale level.

### Measurement invariance

Multi-group invariance by sex and region (UA vs EU) proceeded hierarchically (configural → metric → scalar). Adequacy was judged by ΔCFI ≤ .010 and ΔRMSEA ≤ .015. Where full scalar invariance was untenable, partial scalar invariance was accepted with theory-guided parameter freeing.

### Structural model

We estimated direct paths from Interoceptive Sensibility, Metacognitive Beliefs, and BIS/BAS to IPF (latent), and an indirect path via FSS-9 total. ARS-30 total was evaluated as a moderator (latent interaction or median-split multi-group), with age and sex included as covariates on primary paths.

### Estimation and robustness

Given continuous indicators and potential non-normality, we used robust maximum likelihood (MLR). Indirect effects were evaluated using 5,000-draw bootstrap percentile confidence intervals. Model fit was summarised with χ^2^ (interpreted cautiously), CFI/TLI, RMSEA (90% CI), and SRMR. Sensitivity analyses compared the ESEM solution with (i) a more restrictive CFA and (ii) a reduced MCQ model centred on a Threat/Uncontrollability core index.

### Reporting conventions

To avoid interpretational drift, we reported MCQ-30, BIS/BAS, MAIA-2 as subscale means; FSS-9 as total; IPF as a latent from domain parcels; ARS-30 as total. Standardised coefficients (β) with 95% confidence intervals are presented; two-tailed tests used α = .05. **Aim**. The study aimed to deliver an empirically grounded account of how interoceptive sensibility (MAIA-2), metacognitive beliefs (MCQ-30), and approach–avoidance tendencies (BIS/BAS) jointly shape fatigue (FSS-9) and psychosocial impairment (IPF, coded such that higher scores indicate greater impairment) in a UA/EU higher-education cohort. Specifically, we sought to (i) establish a defensible measurement structure via ESEM and demonstrate measurement invariance by sex and region; (ii) test a structural model in which fatigue carries part of the burden from metacognitive threat and interoceptive sensibility to functioning; and (iii) examine whether academic resilience (ARS-30) buffers these relations. A secondary objective was to articulate behavioural/computational predictions that could be tested in a subsequent post-exertional worsening (PEW) paradigm (Stephan et al., 2016; Greenhouse-Tucknott et al., 2021; Matthews et al., 2023; McMorris, 2020).

### Working hypotheses

We hypothesised that stronger metacognitive threat/uncontrollability and need-to-controlwould be associated with higher fatigue and greater impairment, whereas greater interoceptive sensibility would be associated with lower fatigue and better functioning; approach– avoidance tendencies were expected to show at most modest positive links with fatigue. We further hypothesised partial mediation whereby fatigue transmits a substantive portion of metacognitive and interoceptive effects to impairment, alongside residual direct effects (complementary mediation). We expected academic resilience to attenuate the metacognitive threat → fatigue (and, secondarily, threat → impairment) pathways. Finally, we anticipated measurement invariance across sex and region (at least metric, with partial scalar if required), and we specified that any regional mean differences would not alter the direction or magnitude of core structural paths. For the planned PEW extension, we predicted that exertional challenge would transiently increase fatigue and strengthen the metacognitive-to-fatigue link, a pattern to be modelled trial-wise in future work.

**Table 1.** Measures, scaling, interpretation, and roles in the models (UA/EU)

| Instrument | Format / Range | “Higher score” interpretation | Role in models |
| --- | --- | --- | --- |
| MAIA-2 (UA/EN) | Subscales, item-means | Greater interoceptive sensibility/skills | Indicators of *Interoceptive Sensibility*(ESEM) |
| MCQ-30 | 5 subscales, item-means (1–4) | Stronger metacognitive beliefs (incl. Threat/Uncontrollability) | Indicators of *Metacognitive Beliefs*(ESEM); key predictor |
| BIS/BAS | BIS; BAS-Drive/Fun/Reward, subscale means | Stronger avoidance/approach tendencies | Indicators/observed predictors of *Approach–Avoidance* |
| FSS-9 | Total 9–63 | More severe fatigue | Mediator (Threat → FSS → IPF); severity descriptor |
| IPF | 4 domains, latent | Greater impairment in functioning | Primary outcome (latent criterion) |
| ARS-30 | Total | Higher academic resilience | Moderator of Threat → FSS/IPF (with sensitivity by subscales) |

## Results

The final sample comprised N = 300 students (UA = 151; EU = 149), aged M = 22.8, SD = 3.5; 52% identified as women. Descriptively, fatigue was moderate (FSS-9: M = 30.8, SD = 12.1), and IPF domain scores indicated greater disruption in Work/Study and Social/Friendships than in Self-Care or Family/Intimate (work/study impairment M ≈ 41 on a 0–100 scale; other domains M ≈ 25–34). MAIA-2 subscales clustered in the mid-range (means 2.6–3.1 on 0–5), MCQ-30 subscales centred near 2.2–2.7 (1–4), and BIS/BAS subscales showed the expected spread (BIS ≈ 2.9; BAS-Reward ≈ 3.3).

A three-factor ESEM (Interoceptive Sensibility, Metacognitive Beliefs, Approach–Avoidance) fit the data well: χ^2^(132) = 231.5, p < .001; CFI = .962; TLI = .949; RMSEA = .043, 90% CI [.035, .051]; SRMR = .036. Primary loadings were moderate–strong (MAIA: .55–.82; MCQ: .45–.78; BIS/BAS: .49–.71), with cross-loadings |λ| ≤ .18. Reliability (McDonald’s ω) ranged .76–.88 across domains.

Configural and metric invariance held by sex and by region (UA vs EU) with ΔCFI ≤ .003, ΔRMSEA ≤ .002. Partial scalar invariance was established after freeing two intercepts (MAIA Body Listening; MCQ Cognitive Self-Consciousness) across regions: ΔCFI = .009, ΔRMSEA = .004—adequate for comparing structural paths.

With IPF coded such that higher scores indicate greater impairment, fatigue (FSS-9) showed a robust positiveassociation with IPF (β = .45, 95% CI [.34, .56], p < .001). Metacognitive Beliefs (dominated by Threat/Uncontrollability and Need to Control Thoughts) predicted more fatigue (β = .41 [.29, .52], p < .001) and greater impairment (β = .24 [.10, .38], p = .001). Interoceptive Sensibility predicted less fatigue (β = −.22 [−.33, −.10], p < .001) and better functioning (β = −.19 [−.31, −.07], p = .002). The Approach– Avoidance factor was weakly positive for fatigue (β = .15 [.04, .26], p = .007) and n.s. for IPF (β = .08 [−.03, .18], p = .15). Explained variance was R^2^_FSS = .38and R^2^_IPF = .54.

### Indirect effects (fatigue as mediator)

There was partial mediation of metacognitive influences on functioning via fatigue: Metacognitive Beliefs → FSS → IPF β_indirect = .18 [.10, .27], p < .001. Interoceptive Sensibility showed a protective indirect effect: β_indirect = −.10 [−.17, −.04], p = .001. Direct paths to IPF remained significant, consistent with complementary mediation.

### Moderation by academic resilience (ARS-30)

Academic resilience buffered the link between metacognitive threat and fatigue (latent interaction): β_interaction = −.12 [−.22, −.02], p = .018. Johnson– Neyman probing indicated that the Metacognitive→Fatigue path became non-significant for ARS-30 ≥ +1.2 SD above the mean.

### Group comparisons and sensitivity

EU participants reported slightly lower fatigue than UA (FSS: 29.3 vs 32.2, d = 0.24, p = .041) and marginally better work/study functioning; however, all structural paths were invariant across region and sex. ESEM outperformed a restrictive CFA (ΔBIC = +62), and a reduced MCQ model centred solely on Threat/Uncontrollability degraded fit (ΔCFI = −.024) without altering the direction of key effects.

**Table 2.** Key structural paths, indirect effects, and model fit.

| Effect (standardised $\beta$) | Estimate | 95% CI | p |
| --- | --- | --- | --- |
| Metacognitive Beliefs → FSS | .41 | [.29, .52] | <.001 |
| Interoceptive Sensibility → FSS | -.22 | [-.33, -.10] | <.001 |
| Approach–Avoidance → FSS | .15 | [.04, .26] | .007 |
| FSS → IPF | .45 | [.34, .56] | <.001 |
| Metacognitive Beliefs → IPF (direct) | .24 | [.10, .38] | .001 |
| Interoceptive Sensibility → IPF (direct) | -.19 | [-.31, -.07] | .002 |
| **Indirect:** Metacognitive → FSS → IPF | **.18** | [.10, .27] | <.001 |
| **Indirect:** Interoceptive → FSS → IPF | **-.10** | [-.17, -.04] | .001 |
| ARS-30 × Metacognitive → FSS | -.12 | [-.22, -.02] | .018 |
| **ESEM fit:** $\chi^2(132)=231.5$, CFI=.962, TLI=.949, RMSEA=.043 [.035,.051], SRMR=.036 | | | |

## Discussion

The problem here to be studied is whether individual differences in interoceptive processing and metacognitive appraisal plausibly constitute proximal mechanisms of everyday impairment via their influence on fatigue. Our findings—linking stronger metacognitive threat/uncontrollability beliefs to higher fatigue and poorer psychosocial functioning, with interoceptive sensibility showing a protective pattern—sit squarely within a predictive-processing account of fatigue. In that framework, persistent mismatch between prior expectations and ascending bodily evidence (prediction error) degrades confidence in internal control models; fatigue is then the conscious, metacognitive signal that the expected costs of action outweigh its benefits (Stephan et al., 2016; Greenhouse-Tucknott et al., 2021). The observed indirect pathway from metacognitive threat to impairment via fatigue aligns with this logic: beliefs that one’s internal states are dangerous or uncontrollable would amplify anticipated error and down-weight action policies, thereby worsening day-to-day functioning.

Another way of looking at this question is to embed predictive coding within a cost–benefit controller. Convergent evidence places anterior insula and dorsal anterior cingulate at the interface of interoceptive evidence, error monitoring and effort valuation, with fronto-striatal catecholaminergic systems energising goal pursuit (McMorris et al., 2018; McMorris, 2020; Kok, 2022). Our pattern—metacognitive threat → fatigue; fatigue → impairment; modest approach/avoidance contributions; resilience buffering—maps onto this architecture and echoes task-fMRI work showing overlap between ACC regions recruited by error processing and those correlating with on-task fatigue (Wylie et al., 2017). Computational evidence that momentary fatigue updates as a function of exerted effort and error, and then prospectively alters effort choices, offers a mechanistic bridge from our cross-sectional paths to within-person dynamics (Matthews et al., 2023).

We must therefore hold that “interoception” is not a monolith. While our study indexed interoceptive sensibility (MAIA-2) and found it beneficial, clinical investigations dissect facets with finer grain. In fibromyalgia and ME/CFS, patients can show intact accuracy but elevated “interoceptive prediction error”—a discrepancy between subjective sensibility and objective accuracy— closely tracking fatigue and pain (Sharp et al., 2021). In multiple sclerosis, lower interoceptive insight and white-matter dysconnectivity together increase the odds of cognitive fatigue (Danciut et al., 2024), and questionnaire-based interoceptive awareness and sleep quality predict fatigue out-of-sample (Rouault et al., 2023). Neuroimaging further points to hyperconnectivity among interoceptive hubs with relative disconnection of reward circuitry during fatigue states (Campo et al., 2019; Chen et al., 2020). Our results converge at the construct level but also reveal a limitation: without behavioural accuracy tasks we cannot compute individual-level prediction-error or insight indices. Accordingly, the next empirical step is to add heartbeat-based accuracy and confidence tasks to derive interoceptive insight and an explicit interoceptive prediction-error metric within the same structural framework.

The question therefore naturally arises as to why metacognitive threat beliefs exert such leverage. Teodoro et al. (2018) synthesised evidence across functional neurological disorders, fibromyalgia and CFS to argue that excessive interoceptive monitoring and heightened perception of effort draw attentional resources inward, slowing processing and increasing the felt cost of routine actions. Greenhouse-Tucknott et al. (2021) extend this to a unified model of exertional and pathological fatigue: persistent detection of interoceptive mismatch undermines confidence in control predictions. Our moderation analysis suggests that academic resilience—arguably a proxy for stable control beliefs and adaptive appraisal—attenuates the metacognitive threat→fatigue pathway, which is consistent with the idea that strengthening self-efficacy can recalibrate priors about controllability (Stephan et al., 2016) and with recent proposals placing self-efficacy and affect at the heart of allostatic inference (Krupnik, 2024).

Post-exertional worsening (PEW/PEM) offers a natural perturbation for testing these claims. Learning accounts predict that interoceptive and exteroceptive cues associated with effort can acquire maladaptive predictive value for “feeling tired,” fostering avoidance and generalisation that entrench disability (Lenaert et al., 2018). Predictive-processing models likewise anticipate that exertion transiently elevates interoceptive prediction error and shifts cost–benefit priors (Greenhouse-Tucknott et al., 2021), while computational work shows that errors in cognitive tasks produce momentary spikes in fatigue that alter subsequent decisions (Matthews et al., 2023). Our cross-sectional data cannot adjudicate dynamics, but they furnish the measurement scaffold for a pre-registered PEW paradigm combining MAIA-2, MCQ-30, BIS/BAS, FSS/IPF, and trial-level fatigue/effort modelling.

Finally it can be observed that our inferences cohere with broader neurobiological reviews situating interoception as the afferent arm of body– brain loops subserving homeostasis and allostasis, with disturbances at multiple levels—from afferent signalling through integration to metacognitive appraisal—implicated in fatigue-laden conditions (Quadt et al., 2018). The doctrine provided by this literature suggests intervention targets that are testable within our measurement–structural pipeline: precision-recalibration of bodily signals (biofeedback; skills that reduce sensibility–accuracy mismatch), metacognitive therapy focused on uncontrollability beliefs, motivational framing to shift cost–benefit appraisals, and resilience-building as a trait-level buffer.

Strengths of the present study include a pre-specified ESEM approach with excellent fit, demonstration of metric and partial scalar invariance across sex and region (UA/EU), harmonised reporting that fixes the direction of the IPF outcome, and theoretically anchored moderation and mediation tests. Limitations are the cross-sectional design, reliance on self-report (without behavioural interoceptive accuracy/insight), a student sample that constrains external validity, and the absence of an exertional challenge. These caveats notwithstanding, the pattern of associations is statistically robust and theoretically consonant with predictive-processing and allostatic self-efficacy accounts.

We must conclude that metacognitive threat/uncontrollability, interoceptive sensibility, and resilience form a tractable triad shaping fatigue and day-to-day functioning. From this we can conclude that a promising research programme would combine longitudinal sampling and PEW manipulations with behavioural measures of interoceptive accuracy and insight, computational modelling of trial-wise fatigue, and targeted interventions that aim to recalibrate priors about bodily control. The aim of such work would not be to supplant biological contributors— neuroinflammation, monoaminergic tone, sleep—but to integrate them as modulators of precision and controllability within an inferential controller. In sum, the present study advances a theoretically coherent and measurement-tight bridge between interoceptive inference, metacognition and impairment, and it delineates practical next steps towards mechanism-based assessment and intervention.

## Data Availability

All data produced in the present study are available upon reasonable request to the authors

## Conclusions

We must conclude that individual differences in metacognitive threat/uncontrollability and interoceptive sensibility form a tractable core of the fatigue–functioning nexus. In a UA/EU higher-education cohort (N=300) we established a defensible ESEM structure with metric—and partially scalar— measurement invariance across sex and region, and a structural pattern whereby fatigue (FSS-9) carried a substantive share of the association from metacognitive threat to psychosocial impairment (IPF coded “higher = worse”), while interoceptive sensibility (MAIA-2) showed both direct and indirect protective associations. Approach–avoidance tendencies (BIS/BAS) contributed modestly, and academic resilience (ARS-30) buffered the link between metacognitive threat and fatigue, consistent with an allostatic self-efficacy account.

From this we can conclude that predictive-processing and cost–benefit control frameworks offer a coherent explanation: persistent expectation– evidence mismatch amplifies perceived costs and undermines confidence in bodily control, with fatigue operating as the metacognitive “gatekeeper” that down-weights goal-directed behaviour. Our results align with this doctrine while adding measurement rigour (harmonised scoring, explicit outcome direction, invariance tests) and a practically useful moderation target— resilience—that attenuates risk without eliminating it.

The idea still needs considerable working out. The cross-sectional, student sample and reliance on self-report limit causal inference and generalisability; the absence of behavioural interoceptive accuracy/insight tasks precluded direct estimation of interoceptive prediction error, and no exertional challenge was applied. Nevertheless, the present architecture is statistically robust and theory-congruent, supplying a stable platform for mechanism-driven extensions.

Finally it can be observed that the immediate next steps are clear: (i) integrate heartbeat-based accuracy and confidence measures to compute insight and prediction-error indices within the same SEM; (ii) implement a post-exertional worsening paradigm with trial-wise modelling of fatigue and effort; and (iii) test targeted interventions—metacognitive reframing of uncontrollability beliefs, resilience-building, and precision-recalibrating interoceptive training/biofeedback—with longitudinal follow-up of IPF. We, thus, arrive at the following observation: calibrating priors about bodily control and refining the precision of interoceptive signals are promising, testable levers for reducing fatigue and improving day-to-day functioning.

## Acknowledgements

*This article was prepared with the assistance of artificial intelligence (AI) technologies, including AI-based translation tools, which were used to improve linguistic clarity and to facilitate cross-language editing. All final interpretations, conclusions, and responsibility for the content remain solely with the authors*.

*The authors acknowledge the use of AI-assisted tools (e.g*., *machine translation and text refinement systems) during the preparation of this manuscript. These tools were applied exclusively for language editing purposes and did not affect the scientific interpretation of the results*.

## Author’s clarifications

For the avoidance of doubt, the points below refine terminology and scope without altering analyses, estimates, or conclusions.

1. Interpretation boundary (cross-sectional design). All structural paths are interpreted as associations rather than causes. We resist causal varnish: indirect paths are described as *indirect associations* within a correlational framework.
2. Terminology and scoring. Higher scores on the interference with daily functioning (IPF) index denote *more*interference (i.e., worse functioning). This directionality applies to all tables and figures.
3. Measurement notes. Latent constructs were estimated with an exploratory–structural equation modelling (ESEM) approach. Cross-loadings were permitted a priori to reduce factor-pure bias. Fit indices and full loading matrices are available on request.
4. Moderation analysis. The resilience moderation was estimated within a latent-variable SEM framework. Results are robust to reasonable rescaling of indicators; illustrative simple-slope plots are provided on request.
5. Multiplicity. Given the number of parameters in SEM, inferences prioritise theory-led paths. Secondary contrasts should be read as descriptive and hypothesis-generating.
6. Power and precision. Precision is reported via confidence intervals. We caution that smaller interaction effects may be under-powered and warrant replication.
7. Language and versions. Where adapted language versions of questionnaires were used, semantic equivalence procedures (translation/back-translation) were applied. Cross-language invariance was not the target of this report and remains a task for future work.
8. Data handling. Missingness was addressed under missing-at-random assumptions using full information maximum likelihood. Quality controls (e.g., attention checks, implausible completion time) were applied prior to modelling.
9. Transparency. A prospective analytic plan informed the modelling sequence. A time-stamped copy and a full record of any deviations from the plan are available from the corresponding author and will be deposited in an open repository upon publication.
10. Ethical safeguards. Participation was voluntary with withdrawal at will; support contacts were provided at debrief. No adverse events were reported.
11. Generalisability. The sample represents a convenience cohort. Extrapolation beyond populations with similar sociodemographic features should be made with caution.
12. Future work. To move from association to mechanism, we plan (i) behavioural assays of interoceptive accuracy and metacognitive insight, and (ii) repeated-measures designs to test within-person change and temporal precedence.

